# A Multi-Center, Controlled Human Infection Study of Influenza A (H1N1)pdm09 in Healthy Adults

**DOI:** 10.1101/2022.09.27.22280383

**Authors:** Justin R. Ortiz, David I. Bernstein, Daniel F. Hoft, Christopher W. Woods, Micah T. McClain, Sharon E. Frey, Rebecca C. Brady, Christopher Bryant, Ashley Wegel, Robert W Frenck, Emmanuel B. Walter, Getahun Abate, Sarah R. Williams, Robert L. Atmar, Wendy A. Keitel, Nadine Rouphael, Mathew J. Memoli, Mamodikoe K. Makhene, Paul C. Roberts, Kathleen M. Neuzil

## Abstract

**Background:** Influenza controlled human infection model (CHIM) studies can advance development of vaccines and therapeutics. Our objective was to evaluate the associations between baseline challenge virus-specific hemagglutination inhibition (HAI) and microneutralization (MN) titers and subsequent symptomatic influenza virus infection.

**Methods:** We enrolled healthy adults aged 18 through 49 years in a multisite CHIM study using influenza A/Bethesda/MM2/H1N1, an A/California/04/2009/H1N1pdm-like virus (NCT04044352). We excluded persons vaccinated against influenza within the previous six months. We collected serial safety labs, serum for HAI and MN, and nasopharyngeal swabs for RT-PCR testing. Analyses used the putative seroprotective titer of ≥40 for HAI and MN. The primary clinical outcome was mild-to-moderate influenza disease (MMID), defined as ≥1 post-challenge positive qualitative RT-PCR test with a qualifying symptom/clinical finding.

**Findings:** Of 76 participants given influenza virus challenge, 54 (71.1%) experienced MMID. Clinical illness post-challenge was generally very mild. MMID attack rates among participants with baseline titers ≥40 by HAI and MN were 64.9% and 67.9%, respectively, while MMID attack rates among participants with baseline titers <40 by HAI and MN were 76.9% and 78.3%, respectively. The estimated odds of developing MMID decreased by 19% (odds ratio=0.81; 95% CI: 0.62, 1.06; p=0.126) for every two-fold increase in baseline HAI. There were no deaths, serious adverse events, or other significant adverse events.

**Interpretation:** In a multi-site influenza CHIM study, we assured the safety of our participants and achieved a 71.1% attack rate of MMID. High baseline HAI and MN were associated with protection from illness.

## INTRODUCTION

Controlled human infection models (CHIMs) of influenza virus have been used since the 1930s to advance understanding of infection natural history, clinical characteristics, and immune responses (1-16). Such studies were critical for advancing the development of influenza antivirals and have helped establish the antibody correlate of protection, hemagglutination inhibition (HAI) titer ≥40, used as the regulatory standard for influenza vaccine evaluation (17-19). CHIMs can efficiently evaluate vaccine efficacy, avoiding the high costs of field trials and the uncertainty of community strain circulation and attack rates (20). While there is a precedent for vaccines to achieve licensure based on a CHIM study (21), it is more likely that they would be used in mid-stage clinical development of influenza vaccines by informing the best vaccine candidates to advance into pivotal trials conducted to obtain regulatory approval (22).

In the early 2000s, influenza virus CHIM studies were halted for several years in the United States due to an adverse event of dilated cardiomyopathy ultimately deemed unlikely caused by the influenza B challenge virus (23, 24). Beginning in 2012, the Laboratory of Infectious Diseases at the National Institute of Allergy and Infectious Diseases (NIAID) re-established influenza virus CHIMs in the United States, conducting influenza A (H1N1)pdm09 and influenza A (H3N2) CHIMs without safety concerns arising (25, 26).

In 2018, NIAID made CHIMs a central component to its strategic plan for the development of universal influenza vaccines, citing the limitations of animal models of influenza virus infection and prevention (22). To address gaps in the knowledge base, NIAID committed to the following activities to advance CHIMs: 1) expanding capacity for conducting CHIMs; 2) optimizing and standardizing available influenza virus challenge strains, disease models, and infection methods; 3) using CHIMs to study influenza virus pathogenesis, immunity, and correlates of protection; 4) evaluating next-generation universal influenza vaccine candidates; and 5) comparing data from CHIMs to data from studies of natural infection (22). In 2019, NIAID selected six Vaccine and Treatment Evaluation Unit (VTEU) sites to design and execute a multicenter influenza virus CHIM trial.

## METHODS

### Study design and procedures

We conducted a CHIM study with influenza A/Bethesda/MM2/H1N1, an A/California/04/2009/H1N1pdm-like virus, among healthy adults aged 18 through 49 years. The trial was conducted at academic research sites in four US cities: Baltimore, Maryland; Cincinnati, Ohio; Durham, North Carolina; and Saint Louis, Missouri. It was designed to assess clinical response, immunological responses, and safety of the influenza A (H1N1)pdm09 viral challenge. Study participants provided informed consent and were screened for good health beginning one month prior to viral challenge. We excluded persons with influenza vaccination within the previous six months, risk factors for influenza complications, and allergies to influenza antivirals or two or more classes of antibacterials. Beginning two days prior to viral challenge, we admitted participants into an inpatient research unit, reviewed their eligibility criteria, acclimated them to the inpatient environment, collected baseline clinical specimens, and initiated regular nasopharyngeal (NP) swab specimen collection to assess for a panel of respiratory viruses. Participants received the (H1N1)pdm09 viral challenge on their third inpatient day (Study Day 1). They were subsequently monitored in the inpatient facility for at least 7 days with regular self-reported symptom assessments, clinical evaluation, safety assessment, and specimen collections. Infection prevention and control measures were implemented according to guidelines from each study site and in adherence with US government recommendations (27).

### Study product

The (H1N1)pdm09 challenge virus was produced under GMP conditions by Charles River Laboratories (PA) from a virus seed stock derived by reverse genetics and produced by NIAID (25). The final product contained approximately 5 × 10 m^6^ Median Tissue Culture Infective Dose (TCID_50_)/mL in sucrose phosphate glutamate solution, and the 2 mL inoculum of virus product contained a total of 1×10^7^ TCID_50_. The challenge virus was susceptible to FDA-approved neuraminidase inhibitors and sequencing did not indicate any mutations that conferred baloxavir marboxil resistance (28, 29). Full genomic sequencing was performed on the virus (Lot MM#001) (25). There were four nucleotide changes from the seed virus identified post-manufacture: two each in the hemagglutinin (HA) and neuraminidase (NA). Three of the 4 changes were non-synonymous.

### Clinical procedures

We screened persons for good health by medical history, physical examination, screening blood tests, urine toxicology, pregnancy testing when appropriate, electrocardiogram, and chest radiograph. Persons with abnormal findings at baseline were ineligible. On Study Day 1, we delivered 1 mL of virus product in each nostril to semi-recumbent participants using a nasal sprayer device (Wolfe-Tory Medical, Salt Lake City, Utah).

All participants had standardized clinical evaluations, including vital signs, oxygen saturation, and physical examination daily while they were in the inpatient setting, as well as during follow-up appointments, if clinically indicated. All participants had a repeat electrocardiogram on Study Day 7.

Safety laboratory tests (including white blood cells, absolute lymphocyte count, hemoglobin, platelets, alanine transaminase, and creatinine) were conducted at screening and on Study Days 2, 4, and 8. Blood draws for immunologic assays were obtained prechallenge and on Study Days 2, 4, 6 and 8.

Influenza and other respiratory viruses were assessed from nasopharyngeal NP swabs upon admission to the inpatient facility and daily post-challenge. We tested these specimens by qualitative RT-PCR assay using a CLIA-certified, multiplex respiratory virus assay (BIOFIRE^®^ FILMARRAY^®^ or Luminex xTAG^®^). Participants with viral infections detected prechallenge were discharged from the unit and discontinued from the study.

Participants reported their symptoms twice daily during their inpatient stay using a standardized, validated symptom diary (FLU-PRO) (30-32). The 32-item instrument assessed presence, severity, and duration of symptoms across six domains (nose, throat, eyes, chest/respiratory, gastrointestinal, and systemic). The FLU-PRO total score is computed as a mean score across all 32 items. Total scores can range from 0 (symptom free) to 4 (very severe symptoms).

Beginning on Study Day 8, participants could leave the inpatient unit if they had: 1) two consecutive negative influenza A qualitative RT-PCR tests; and 2) were afebrile (<38.0°C), had a room air SpO2 ≥95%, and had been clinically and hemodynamically stable for at least 48 hours. All participants with positive influenza tests on Study Day 8 were offered one dose of baloxavir marboxil (a full treatment course). Participants underwent scheduled clinical evaluation, blood draws, NP swabs, and safety laboratory tests on Study Days 15, 29, and 61. A final telephone call occurred on Study Day 91 to assess safety.

### Research laboratory evaluation

Immunologic testing included virus-specific HAI and microneutralization (MN) assays (Southern Research, Birmingham, AL) qualified for A/California/09 H1N1 and described previously (33-36), and a fit-for-purpose neuraminidase inhibition (NAI) assay by Enzyme Linked Lectin Assay (ELLA) (Battelle, West Jefferson, OH) against a reassortant H6N1 strain with HA from A/turkey/Massachusetts/3740/1975, NA from A/California/04/2009, and internal proteins from A/Puerto Rico/8/1934 (37).

### Statistics

We planned a sample size of 80 participants, but the study was not designed to test any specific null hypothesis or reach a pre-determined level of estimation precision. The primary objective was to evaluate the association between baseline HAI antibody titers and the development of mild-to-moderate influenza disease (MMID) post-challenge. Secondary objectives included determining the frequency of serious adverse events (SAEs), assessing viral recovery via quantitative RT-PCR, describing HAI and MN responses post-challenge by infection status, and evaluation of associations between asymptomatic influenza virus infection or symptomatic influenza negative status and pre-existing HAI antibody titers. Exploratory analyses included describing NAI titers and infection post-challenge.

We conducted analyses according to an *a priori* analysis plan. Clinical and immunological analyses were conducted in the modified Intent-to-Treat (mITT) population, consisting of all participants who received the viral challenge and were followed through the inpatient period (through Study Day 8) or at least until the outcome in question was evaluable. Analyses of safety data and symptom data not involving immunological components were conducted in the Safety Population, which included all participants who received the influenza viral challenge.

MMID required two criteria occurring at any time during seven days post-challenge: 1) evidence of influenza virus as detected by qualitative RT-PCR assay from a NP swab; and 2) any one or more of the following: arthralgia, chest tightness, chills, conjunctivitis, coryza, decreased appetite, diarrhea, dry cough, dyspnea/shortness of breath, fatigue/tiredness, fever (>38.0°C), headache, lymphopenia (<1000 cells/mL), myalgia, nasal congestion, nausea, oxygen saturation decrease by ≥3% from baseline, productive cough, rhinorrhea, sinus congestion, sore throat, and sweats (25).

Participants were classified into post-challenge categories: MMID, asymptomatic with at least one influenza A RT-PCR detection, or no influenza A RT-PCR detection. For a secondary analysis to examine an alternate case definition, MMID-2 required at least two positive RT-PCR tests of NP swabs collected post-challenge in addition to the same symptom criteria as MMID.

HAI, MN, and NAI titer values were reported as an inverse dilution with a lower limit of detection of 10. Results not detected at the lowest dilution were imputed as half of the lower limit. Individual responses were computed as the geometric mean of replicate titers. Serum assay results were summarized by timepoint using geometric mean titers (GMTs) and geometric mean fold-rise (GMFR), along with associated 95% confidence intervals (CIs) based on the t-distribution on the log-scale. CIs for geometric mean values were estimated based on the assumption of log-normality. We defined seroconversion for HAI, MN, and NAI as a baseline titer ≥10 and a minimum four-fold rise in post-challenge titer, or a baseline titer <10 and a post-challenge titer ≥40. We defined seroprotection for HAI and MN titers by a titer ≥40, reflecting a putative cut point commonly used to define susceptible vs seroprotective titers (38). We did not define a seroprotective titer for NAI. Exact Clopper-Pearson CIs were calculated for these and other binary endpoints. For statistical comparisons, p values <0.05 were considered statistically significant.

For the primary analysis, the association between baseline HAI antibody titer and MMID (as observed through Study Day 8) was evaluated via univariable logistic regression. The associations of baseline MN and NAI titers with MMID were assessed similarly.

Viral shedding was assessed by evaluating the frequency, timing, magnitude (peak and total viral shedding) and duration overall and by baseline HAI, MN, or NAI. Kaplan-Meier methodology was used to estimate time to detectable infection, with the censoring of early terminations and participants without detectable virus throughout the challenge period. The duration of viral shedding was evaluated from the day of the initial qualitative RT-PCR positive result to the day of the last positive result, regardless of intermittent negative results. Peak and total viral load (VL) were measured by quantitative RT-PCR. Total viral shedding was estimated via the area under the curve (AUC) of viral copies/mL over the challenge period. Mean differences were computed between baseline HAI and MN seroprotection status groups for peak and total viral shedding, along with associated 95% CIs.

Adverse events (AEs) were coded into Medical Dictionary for Regulatory Activities (MedDRA^®^) system organ class (SOC) and preferred terms. The number and percentage of participants reporting at least one unsolicited AE (serious or non-serious) were summarized by SOC and preferred term, along with associated exact 95% CIs. The incidence and frequency of unsolicited AEs were summarized by SOC and preferred term over time and at any point over the course of the study; severity and relatedness were described as well. For incidence calculations, each participant was counted once under the maximum severity or the strongest recorded causal relationship to viral challenge for a given event.

### Ethics

The protocol and informed consent forms were approved by the institutional review boards at the participating VTEUs. Study participants provided informed consent. The study is registered at ClinicalTrials.gov (NCT04044352).

## RESULTS

### Participant demographics and baseline immunologic results

From October 8, 2019, through February 4, 2020, we screened 188 people; 76 received influenza virus challenge (Figure 2). Demographics and baseline influenza antibody titers varied across the four study sites (Table 1 and Supplemental Table 2). The mean age was 33.4 years (range 18-49), 46 (61%) were male, 5 (7%) were Hispanic, and 35 (46%) were Black or African American. Sixty-seven (88%) participants reported not receiving the seasonal influenza vaccine during the previous year (2018-19). Among participants receiving the influenza virus challenge, 75 (99%) completed the inpatient phase and 65 (86%) completed all study visits. The Geometric Mean Titers (GMTs) (95% CIs) prior to challenge for HAI, MN, and NAI were 42.8 (5.0-718.4), 79.7 (5.0-1280.0), and 651.8 (51.0-3412.0), respectively (Table 2). Thirty-nine (51.3%) participants had baseline HAI titers ≥40 and 23 (30.3%) had baseline MN titers ≥40.

**Figure 1.**
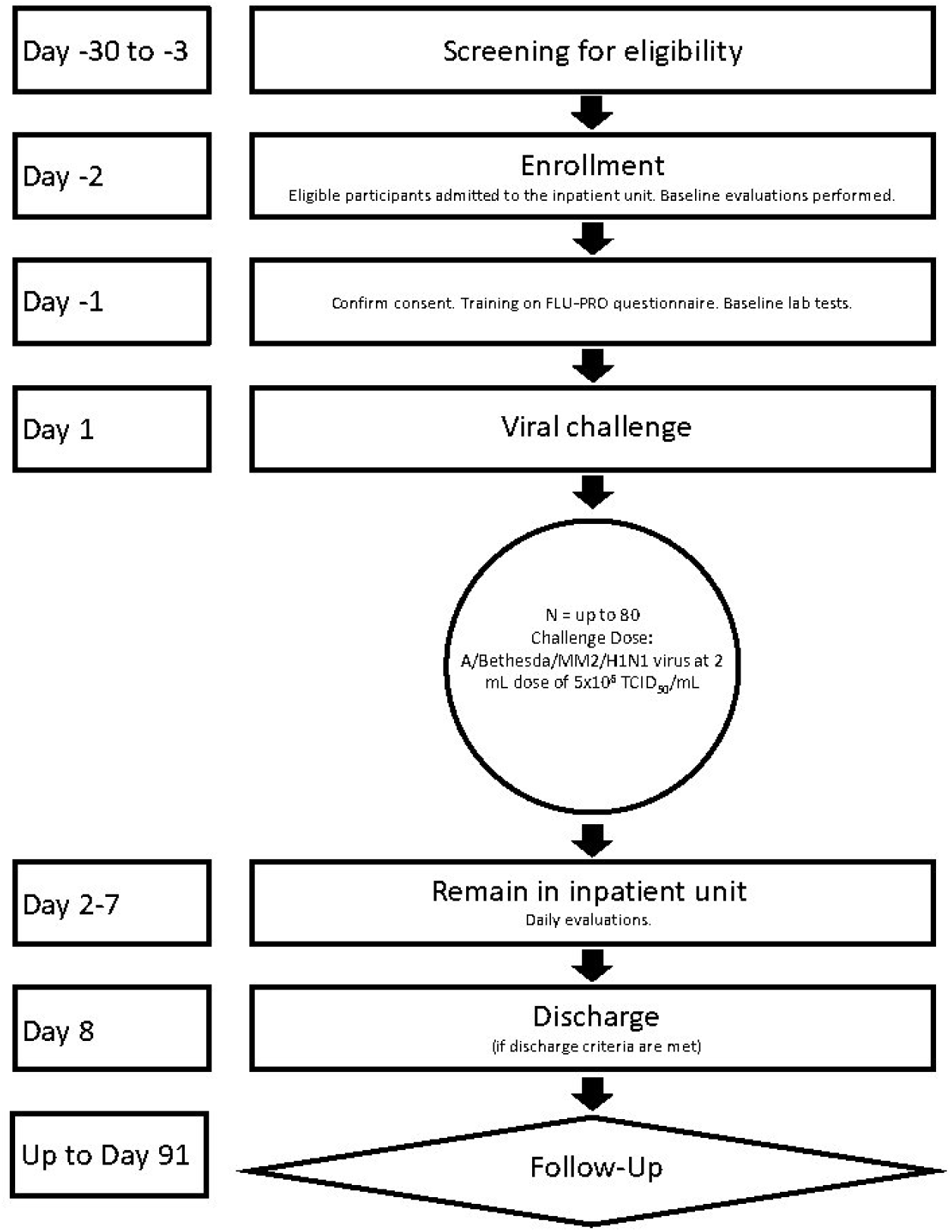
Schematic of study design.

**Table 1.** Participant demographics, baseline antibody status, and post-challenge outcomes.

| Characteristic | Baltimore, Maryland (N=20) | Cincinnati, Ohio (N=24) | Durham, North Carolina (N=15) | St. Louis, Missouri (N=17) | All Participants (N=76) |
| --- | --- | --- | --- | --- | --- |
| Age in years, mean (standard deviation) | 35.0 (7.9) | 32.3 (8.9) | 34.9 (11.8) | 31.7 (17) | 33.4 (9.2) |
| Sex, number (%) |  |  |  |  |  |
| Male | 14 (70.0) | 17 (70.8) | 6 (40.0) | 9 (52.9) | 46 (60.5) |
| Female | 6 (30.0) | 7 (29.2) | 9 (60.0) | 8 (47.1) | 30 (39.5) |
| Ethnicity, number (%) |  |  |  |  |  |
| Not Hispanic or Latino | 17 (85.0) | 24 (100) | 14 (93.3) | 16 (94.1) | 71 (93.4) |
| Hispanic or Latino | 3 (15.0) | 0 (0.0%) | 1 (6.7) | 1 (5.9) | 5 (6.6) |
| Race/ethnicity, number (%) |  |  |  |  |  |
| Black or African American | 14 (70.0) | 11 (45.8) | 8 (53.3) | 2 (11.8) | 35 (46.1) |
| White | 3 (15.0) | 13 (54.2) | 5 (33.3) | 13 (76.5) | 34 (44.7) |
| Other | 3 (15.0) | 0 (0.0) | 2 (13.3) | 2 (11.8) | 7 (9.2) |
| BMI*, mean (standard deviation) | 29.3 (4.1) | 27.2 (4.1) | 28.8 (3.8) | 25.3 (4.1) | 27.6 (4.3) |
| Prior seasonal influenza vaccination (2018-2019), number (%) |  |  |  |  |  |
| No | 19 (95.0) | 23 (95.8) | 13 (86.7) | 12 (70.6) | 67 (88.2) |
| Yes | 1 (5.0) | 1 (4.2) | 2 (13.3) | 5 (29.4) | 9 (11.8) |
| Baseline hemagglutination inhibition titer $\geq 40$, number (%)<sup>†</sup> | | | | | |
| No | 10 (50.0) | 13 (54.2) | 11 (73.3) | 5 (29.4) | 39 (51.3) |
| Yes | 10 (50.0) | 11 (44.8) | 4 (26.7) | 12 (70.6) | 37 (48.7) |
| Baseline microneutralization titer $\geq 40$, number (%)<sup>†</sup> | | | | | |
| No | 8 (40.0) | 9 (37.5) | 2 (13.3) | 4 (23.5) | 23 (30.3) |
| Yes | 12 (60.0) | 15 (62.5) | 13 (86.7) | 13 (76.5) | 53 (69.7) |
| Clinical outcomes, number (%) |  |  |  |  |  |
| MMID<sup>‡</sup> | 14 (70.0) | 20 (83.3) | 10 (66.7) | 10 (58.8) | 54 (71.1) |
| MMID-2<sup>§</sup> | 9 (45.0) | 13 (54.2) | 8 (53.3) | 8 (47.1) | 38 (50.0) |
| Asymptomatic with at least one influenza detection | 3 (15.0) | 1 (4.2) | 4 (26.7) | 0 (0.0) | 8 (10.5) |
| No influenza detections | 3 (15.0) | 3 (12.5) | 1 (6.7) | 7 (41.2) | 14 (18.4) |

\*BMI=body mass index
<sup>†</sup>We defined seroprotection for hemagglutination inhibition and microneutralization titers by a titer $\geq 40$ , reflecting a putative cut point commonly used to define susceptible vs seroprotective titers. We did not define a seroprotective titer for NAI.
<sup>#</sup>MMID= mild-to-moderate influenza disease including at least 1 influenza A detection (primary clinical endpoint)
<sup>§</sup>MMID-2= mild-to-moderate influenza disease including at least 2 influenza A detections (secondary clinical endpoint)

**Table 2.** Baseline hemagglutination inhibition, microneutralization, and neuraminidase inhibition antibodies and post-challenge outcomes.

|  | RT-PCR* negative<br>(none positive) | RT-PCR* positive<br>(one or more) | RT-PCR* positive<br>(two or more) | MMID<sup>†</sup> | MMID-2<sup>‡</sup> | RT-PCR* positive<br>asymptomatic |
| --- | --- | --- | --- | --- | --- | --- |
| n<sup>§</sup> | 14 | 62 | 43 | 54 | 38 | 8 |
| **Hemagglutination inhibition** |  |  |  |  |  |  |
| GMT<sup>¶</sup> (95% CI) | 117.9 (61.3,227.0) | 34.0 (24.7,47.0) | 26.3 (18.0,38.6) | 36.8 (26.1, 52.0) | 27.6 (18.4,41.4) | 20.0 (6.9, 58.0) |
| % with titer ≥40<br>(95% CI)<sup>¶¶</sup> | 78.6 (49.2,95.3) | 41.9 (29.5,55.2) | 32.6 (19.1,48.5) | 44.4 (30.9,58.6) | 34.2 (19.6,51.4) | 25 (3.2, 65.1) |
| **Microneutralization** |  |  |  |  |  |  |
| GMT<sup>¶</sup> (95% CI) | 276.7 (124.8, 613.1) | 60.1 (42.3, 85.4) | 45.2 (30.0, 68.2) | 62.9 (43.1,91.9) | 45.1 (29.3, 69.5) | 44.3 (13.8, 142.6) |
| % with titer ≥40<br>(95% CI)<sup>¶¶</sup> | 92.9 (66.1, 99.8) | 64.5 (51.3, 76.3) | 60.5 (44.4, 75) | 66.7 (52.5, 78.9) | 60.5 (43.4, 76) | 50 (15.7, 84.3) |
| **Neuraminidase inhibition** |  |  |  |  |  |  |
| GMT<sup>¶</sup> (95% CI) | 844.2<br>(508.1,1402.7) | 614.8 (458.4,824.4) | 565.4 (396.0,807.2) | 584.6 (424.7,804.7) | 506.1 (345.4,741.6) | 863.3<br>(358.3,2080.3) |

\*RT-PCR=qualitative reverse transcription polymerase chain reaction
†MMID= mild-to-moderate influenza disease including at least 1 influenza A detection (primary clinical endpoint)
‡MMID-2= mild-to-moderate influenza disease including at least 2 influenza A detections (secondary clinical endpoint)
§n = Number of participants in the Modified Intent-to-Treat population with available results
¶GMT = Geometric Mean Titer
¶¶We defined seroprotection for hemagglutination inhibition and microneutralization titers by a titer ≥40, reflecting a putative cut point commonly used to define susceptible vs seroprotective titers. We did not define a seroprotective titer for NAI.

**Figure 2.**
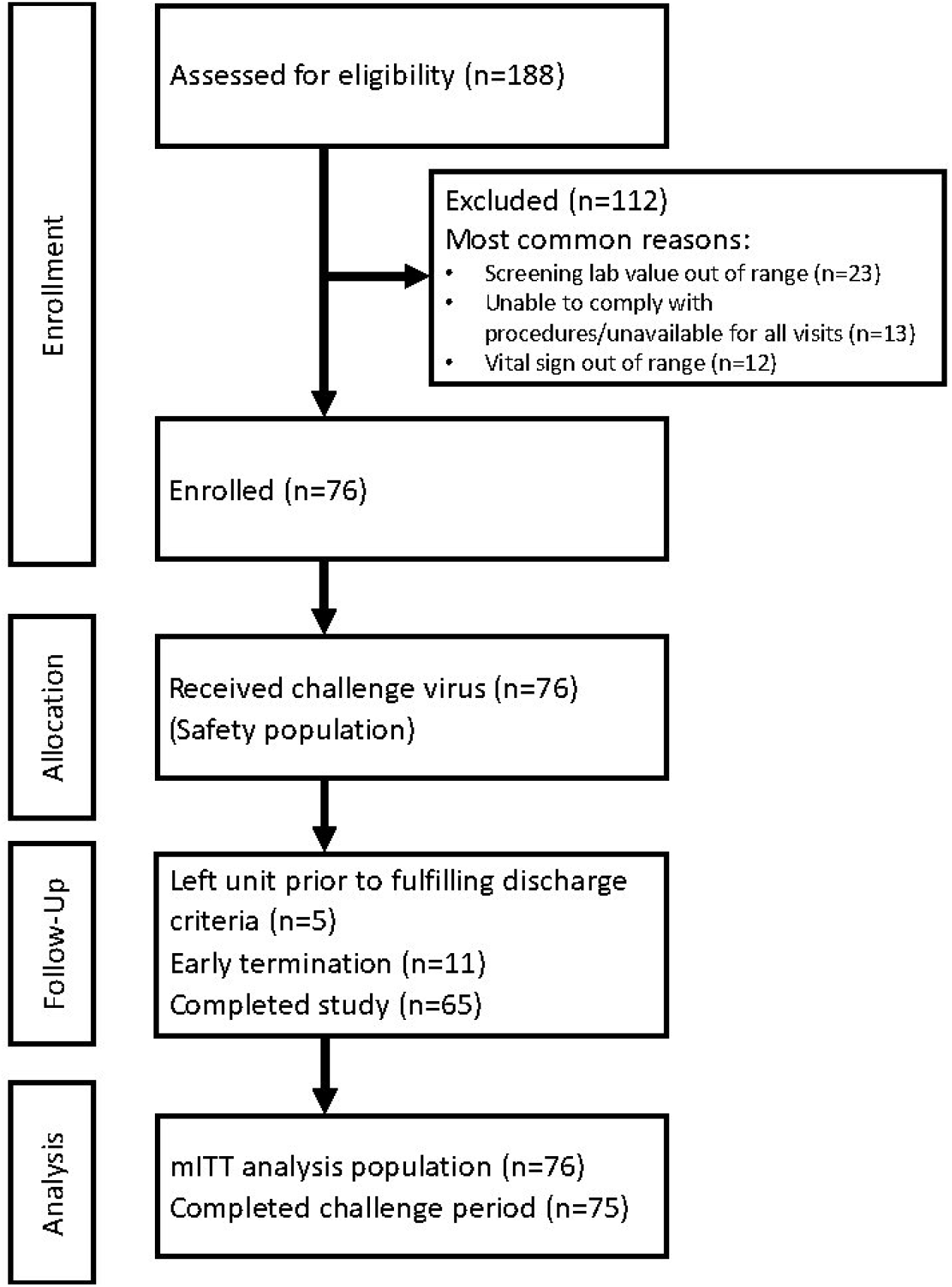
Consort flow diagram. Footnotes: mITT=Modified Intent-to-Treat

### Influenza virus detection

Among participants receiving the influenza virus challenge, 62 (81.6%) had at least one influenza A virus detection and 43 (56.6%) had at least two influenza A detections through Study Day 8 (Table 2). The frequency of influenza A detections peaked on Study Day 2 and decreased rapidly (Supplemental Figure 1). Sixty (78.9%) participants had an initial influenza A detection on Study Day 2, and two (2.6%) had an initial influenza A detection on Study Day 3. By Study Day 8, six (7.9%) participants still had an influenza A detection and were treated with baloxavir. One participant (1.3%) had influenza A detections through Study Day 13, and one (1.3%) left the challenge unit prior to Study Day 8 after several negative influenza A tests.

Baseline HAI and MN were negatively associated with influenza A detection post-challenge (Table 2). Participants without any influenza A detection (n=14) had baseline HAI GMT of 117.9 (95% CI: 61.3, 227.0), MN GMT of 276.7 (95% CI: 124.8, 613.1), and NAI GMT of 844.2 (95% CI: 508.1, 1402.7). Participants with an influenza A detection (n=62) had baseline HAI GMT of 34.0 (95% CI: 24.7, 47.0), MN GMT of 60.1 (95% CI: 42.3, 85.4), and NAI GMT of 614.8 (95% CI: 458.4, 824.4). The proportions of participants with baseline titers ≥40 by HAI and MN among those with any influenza A detection were 41.9% (95% CI: 29.5%, 55.2%) and 64.5% (95% CI: 51.3%, 76.3%), respectively, while the proportions of participants with baseline titers ≥40 by HAI and MN among those without influenza A detections was 78.6% (95% CI: 49.2%, 95.3%) and 92.9% (95% CI: 66.1%, 99.8%). Using the more stringent definition for viral shedding of ≥2 days showed an even greater difference between these groups (Table 2).

The mean duration of viral shedding by qualitative RT-PCR for the 61 participants with at least one influenza A detection who were followed for the entire challenge period was 3.1 days (95% CI: 2.5, 3.8). The duration tended to be higher in participants with baseline HAI titer <40 (n=35), with a mean of 3.8 days (95% CI: 2.9, 4.7), compared to participants with baseline HAI titer ≥40 (n=26), with a mean of 2.3 days (95% CI: 1.6, 3.0), with overlapping 95% CIs. Participants with baseline MN titer <40 (n=23) shed virus longer on average [mean of 3.9 days (95% CI: 2.6, 5.2)] than participants with baseline MN titer ≥40 (n=39) [mean of 2.7 days (95% CI: 2.1, 3.3)], also with overlapping 95% CIs. The frequency of influenza A detection by study day was consistently higher among those with baseline titer <40 for both HAI and MN.

Peak and total viral load by quantitative RT-PCR were both significantly lower in participants with baseline HAI titer ≥40 vs. <40 (difference in mean peak VL of 2.6 log-10 copies/mL, 95% CI: 0.8, 4.4; difference in mean AUC of 2.8 log-10 copies/mL per day observed, 95% CI: 1.0, 4.6). The differences between MN seroprotection groups were slightly smaller and did not meet statistical significance.

### Influenza illness

Fifty-four (71.1%) participants met the MMID case definition, while 38 (50.0%) met the MMID-2 case definition (Table 1. **Participant demographics, baseline antibody status, and post-challenge outcomes**). For the primary analysis of the association of baseline HAI titer and the subsequent development of MMID, univariable logistic regression estimated 19% decrease in the odds of MMID for every two-fold increase in baseline HAI titer (i.e., one unit increase in log-2 titer) [OR: 0.81 (95% CI 0.62, 1.06); p=0.126] (Table 3). In a secondary analysis, there was a stronger association between baseline HAI titer and development of MMID-2 – an estimated 32% decrease in the odds of MMID-2 for every two-fold increase in baseline HAI titer [OR: 0.68 (95% CI 0.52, 0.89); p=0.006]. Univariable logistic regression estimated a 23% decrease in the odds of MMID for every fold increase in baseline MN titer [OR: 0.77 (95% CI 0.60, 0.98); p=0.035] and an estimated 33% decrease in the odds of MMID-2 for every fold increase in baseline MN titer [OR: 0.67 (95% CI 0.52, 0.86); p=0.002] (Table 3). There was a non-significant association between baseline NAI titer and development of MMID [OR: 0.80 (95% CI 0.57, 1.11); p=0.183], but there was evidence of an association between baseline NAI titer and development of MMID-2 [OR: 0.74 (95% CI 0.55, 1.00); p=0.050] (Table 3).

**Table 3.** Univariable logistic regression models* evaluating the relationship of baseline antibodies with MMID^†^ and MMID-2^‡^.

|  | Model parameter | Parameter estimate | Standard Error | p-value | Odds Ratio | 95% CI |
| --- | --- | --- | --- | --- | --- | --- |
| **MMID** |  |  |  |  |  |  |
| Hemagglutination inhibition<sup>§</sup> | Intercept | 2.06 | 0.82 | 0.012 | - | - |
|  | Baseline log-2 Titer<sup>□</sup> | -0.21 | 0.14 | 0.126 | 0.81 | 0.62, 1.06 |
| Microneutralization | Intercept | 2.65 | 0.90 | 0.003 | - | - |
|  | Baseline log-2 Titer<sup>□</sup> | -0.27 | 0.13 | 0.035 | 0.77 | 0.60, 0.98 |
| Neuraminidase inhibition | Intercept | 3.02 | 1.64 | 0.065 | - | - |
|  | Baseline log-2 Titer<sup>□</sup> | -0.22 | 0.17 | 0.183 | 0.80 | 0.57, 1.11 |
| **MMID-2** |  |  |  |  |  |  |
| Hemagglutination inhibition | Intercept | 2.05 | 0.78 | 0.008 | - | - |
|  | Baseline log-2 Titer<sup>□</sup> | -0.38 | 0.14 | 0.006 | 0.68 | 0.52, 0.89 |
| Microneutralization | Intercept | 2.56 | 0.85 | 0.003 | - | - |
|  | Baseline log-2 Titer<sup>□</sup> | -0.41 | 0.13 | 0.002 | 0.67 | 0.52, 0.86 |
| Neuraminidase inhibition | Intercept | 2.80 | 1.45 | 0.054 | - | - |
|  | Baseline log-2 Titer<sup>□</sup> | -0.30 | 0.15 | 0.050 | 0.74 | 0.55, 1.00 |

\*76 subjects in the Modified Intent-to-Treat population included in the models
†MMID= mild-to-moderate influenza disease including at least 1 influenza A detection (primary clinical endpoint)
‡ MMID-2= mild-to-moderate influenza disease including at least 2 influenza A detections (secondary clinical endpoint)
§primary study analysis
□Baseline log-2 Titer calculated based on subject-specific geometric mean titer values for each assay, then log-2 transformed

Based on a univariable logistic regression, each two-fold increase in baseline MN titer was associated with a 42% [OR: 0.58 (95% CI 0.41, 0.82)] decrease in the odds of becoming RT-PCR positive and symptomatic (i.e., meeting MMID criteria) compared to remaining RT-PCR negative, and a 49% [OR: 0.51 (95% CI 0.31, 0.83)] decrease in odds of being RT-PCR positive and asymptomatic compared to remaining RT-PCR negative.

Predicted probabilities of MMID according to baseline HAI, MN, or NAI from univariable models are in Supplemental Figures 2-4. Kaplan-Meier curves with cumulative infection probabilities by HAI and MN baseline titers ≥40 are in Supplemental Figures 5-6.

Symptom scores were low overall, with Study Day 3 mean scores of 0.25 (95% CI: 0.16, 0.36) among participants with at least one influenza A virus detection and 0.10 (95% CI: 0.06, 0.15) among participants without influenza A virus detection. The most common symptoms were nasal congestion/rhinorrhea, sore throat, and sinus congestion. Only 3 (4.8%) had a post-challenge fever (>38.0° C). Symptom scores by body system/domain peaked on Study Day 3, with generally higher scores observed among persons with any influenza A detection compared to those without influenza A detection (Figure 3), with statistically significant differences on Study Days 3, 5, and 6.

**Figure 3.**
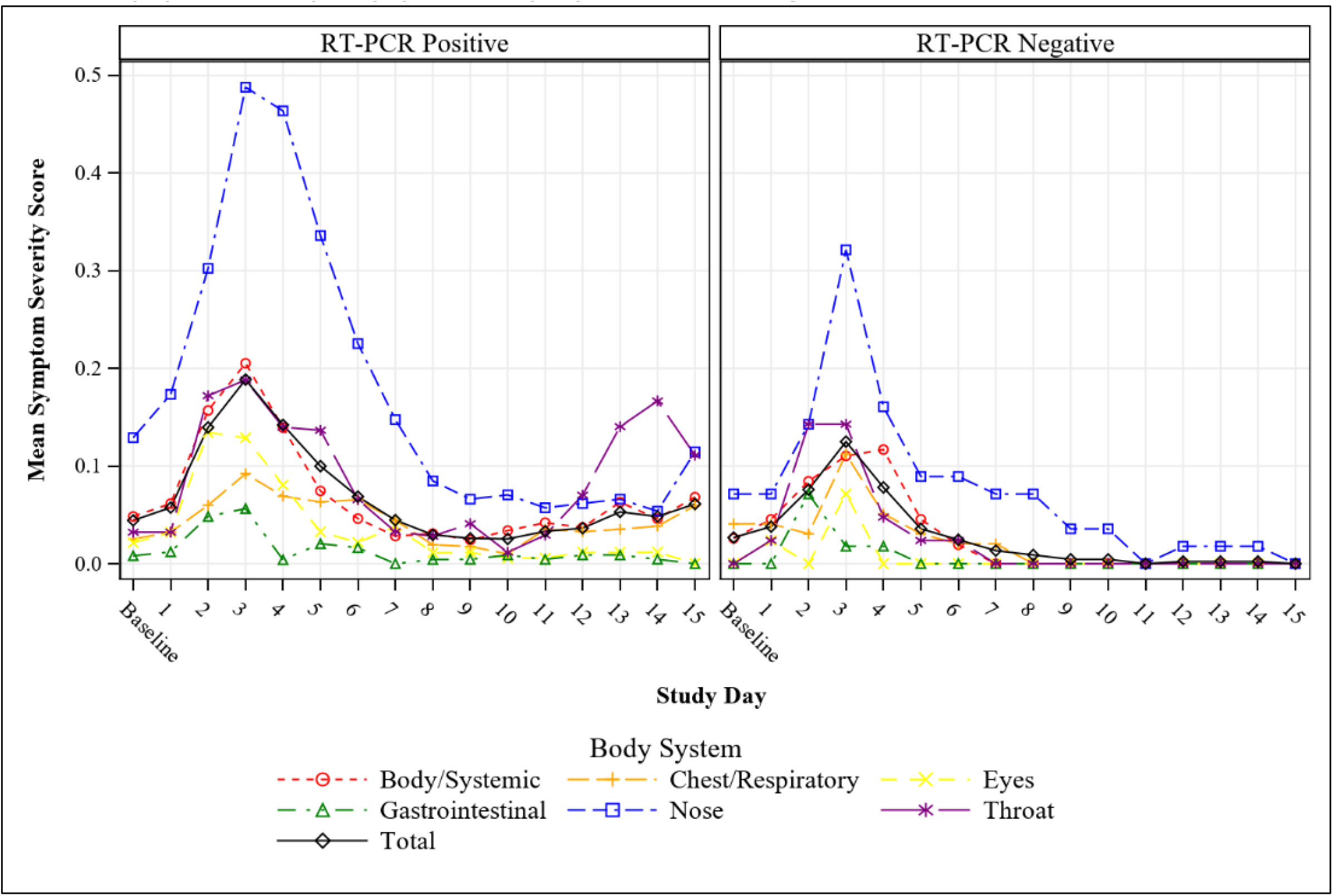
Mean symptom score by body system, study day, and viral shedding status. Footnotes: RT-PCR=qualitative reverse transcription polymerase chain reaction

Participants with higher baseline antibody titers had qualitatively fewer MMID cases when infected than participants with lower titers (Table 2). MMID cases (n=54) had baseline HAI GMT of 36.8 (95% CI: 26.1, 52.0), MN GMT of 62.9 (95% CI: 43.1, 91.9), and NAI GMT of 584.6 (95% CI: 424.7, 804.7). Asymptomatic participants with influenza A detections (n=8) had baseline HAI GMT of 20.0 (95% CI: 6.9, 58.0), MN GMT of 44.3 (95% CI: 13.8, 142.6), and NAI GMT of 863.3 (95% CI: 358.3, 2080.3). Baseline seroprotection by HAI and MN among MMID cases was 44.4% (95% CI: 30.9%, 58.6%) and 66.7% (95% CI: 52.5%, 78.9%). Baseline seroprotection by HAI and MN among asymptomatic participants with influenza A detection was 25.0% (95% CI: 3.2%, 65.1%) and 44.3% (13.8%, 142.6%).

### Immune response to challenge

GMFR from baseline and seroconversion proportions over time (baseline and Day 8, 29 and 61 post-challenge) by post-challenge infection status are in Figure 4, Supplemental Tables 3-5, and Supplemental Figure 7. HAI and MN titers tended to peak at Day 29 and level off through Day 61 for participants with any influenza A detection, while participants without influenza A detection maintained similar levels to baseline. NAI titers also peaked at Day 61 among participants with any influenza A detection. No participants without influenza A detection (n=14) seroconverted for HAI, MN, or NAI. Among MMID cases (n=54) and asymptomatic participants with at least one influenza A RT-PCR detection (n=8), the estimated probability of seroconversion by HAI was similar at Day 8 [3.8% (95% CI: 0.5%, 13.0%) vs. 0.0% (0.0%, 36.9%), respectively] and at Day 29 [28.6% (3.7%, 71.0%) vs. 28.3% (16.0%, 43.5%), respectively], but it was qualitatively higher in the small group of asymptomatic participants than MMID cases at Day 61 [33.3% (95% CI 4.3%, 77.7%) vs. 18.6% (95% CI 8.4%, 33.4%), respectively; (NS)]. The same pattern was observed in seroconversion for MN and NAI over time. For NAI, participants without influenza A detection and asymptomatic participants with at least one influenza A detection tended to have higher titers than MMID cases at each study timepoint, although this pattern was less distinct than for the other assays.

**Figure 4.**
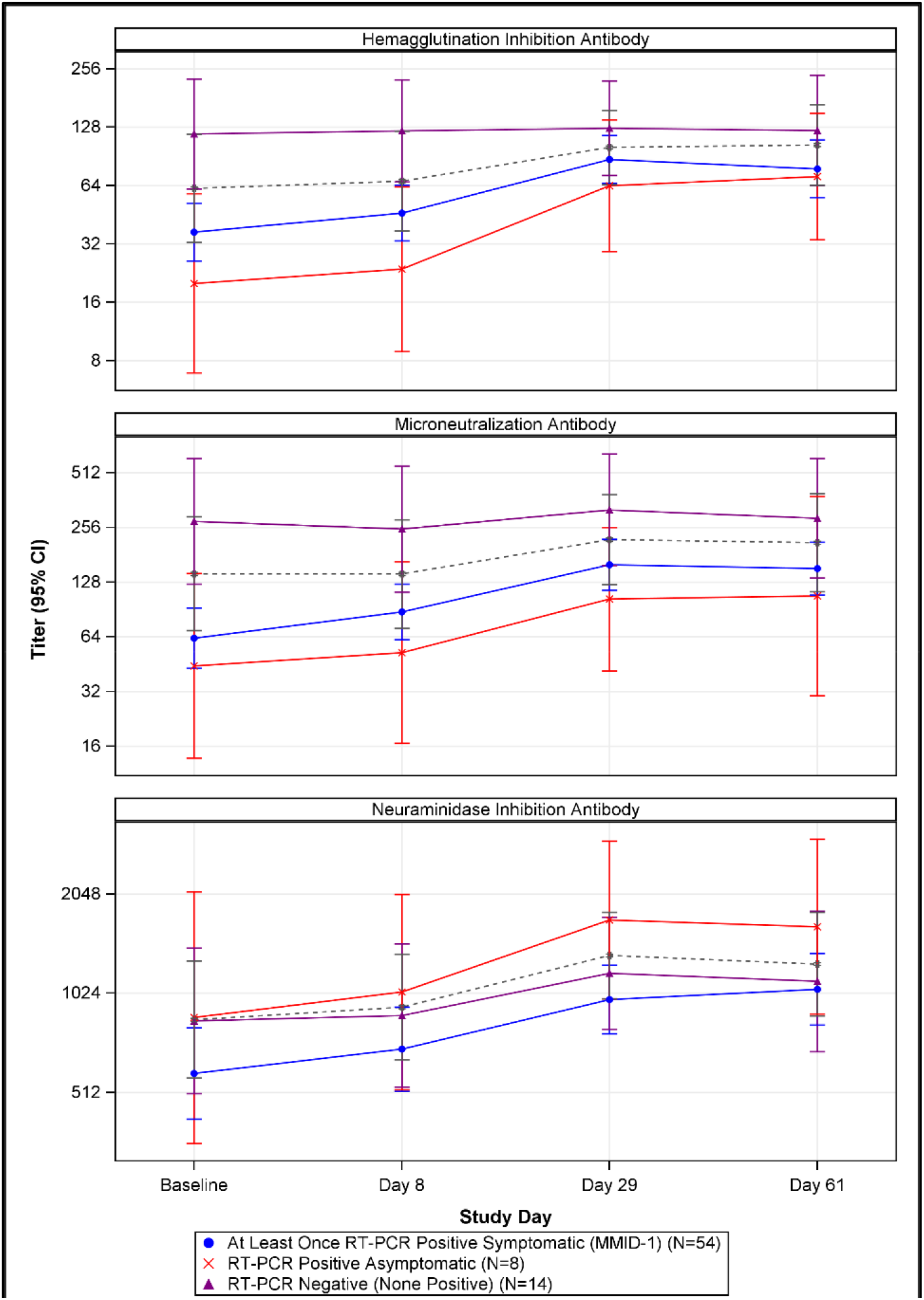
Hemagglutination inhibition, microneutralization, and neuraminidase inhibition antibody geometric mean titers, by infection status and study day. Footnotes: • RT-PCR=qualitative reverse transcription polymerase chain reaction • MMID defined as: One or more PCR+ tests and meeting the symptom threshold at any point during the challenge period • The gray dashed line displays the trajectory of the combined RT-PCR positive asymptomatic and RT-PCR negative groups (i.e., participants who did not meet the definition of MMID).

### Safety

Of the 76 participants in the Safety Population, 46 (60.5%) experienced an unsolicited AE of any attribution, including 26 participants (34.2%) who experienced at least one challenge-related unsolicited AE (Supplemental Table 6). Challenge-related unsolicited AEs were mostly mild in severity (24/76, 31.6%), with two participants (2/76, 2.6%) experiencing challenge-related unsolicited AEs of moderate severity (ear pain and pyrexia) and no severe unsolicited AEs reported. The most common unsolicited AE was lymphadenopathy, occurring in 10 (13.2%) participants; all were mild in severity. No non-influenza respiratory viruses were detected post-challenge. There were no post-challenge ECG changes or cardiac abnormalities. There were no deaths or other SAEs, and no participants discontinued due to AEs. Laboratory abnormalities graded as AEs were all mild in severity (hematology: 13/76 [17.1%]; chemistry: 7/76 [9.2%]).

## DISCUSSION

In this multicenter study, intranasal administration of 1×10^7^TCID_50_ influenza A (H1N1)pdm09 virus resulted in overall attack rates of 71%, 50%, and 10%, respectively of MMID, MMID-2, and asymptomatic influenza virus infection. Clinical illness post-challenge was generally very mild. Participants were diverse, unvaccinated, and almost evenly divided by baseline HAI titers <40 and ≥40. The HAI GMT at baseline was higher in those individuals who did not subsequently develop MMID than in those who did, although not all these differences were statistically significant. Likewise, higher baseline HAI was generally associated with a reduction in overall viral detection and mean duration of viral detection. Corresponding results for analysis of baseline MN titers may have been more discerning.

Our study was modelled on an influenza A (H1N1)pdm09 CHIM previously reported by NIAID researchers with a 68% attack rate of MMID using 1×10^7^ TCID_50_ dose (39). Similarly, we found that baseline HAI titer was associated with influenza outcomes, though the statistical significance of these associations was variable. The NIAID CHIM study reported that baseline NAI titer was more strongly correlated with clinical protection post-challenge than HAI (39), while we observed a weaker association between baseline NAI titer and influenza outcomes. A key difference between the NIAID study and ours was that the former excluded persons with challenge virus-specific HAI titers >40 assessed at approximately eight weeks prior to challenge. Some participants likely had subsequent, pre-challenge influenza exposures, as HAI titers were higher by the time of inpatient admission (39). Nevertheless, excluding participants with baseline HAI titers ≥40 likely limits the ability of a study to evaluate associations of baseline HAI titers and infection or clinical influenza outcomes. Other immune responses that correlate with HAI may also be similarly affected.

Symptom severity in our study was consistent with previous influenza CHIMs (30), including the NAID study with the same virus (25). On Study Day 3, we measured peak average symptom score of 0.25 among participants with influenza A detections and 0.10 among participants without influenza A detection. This is equivalent to participants with influenza A detection reporting eight mild symptoms on the 32-item FLU-PRO instrument and participants without influenza A detection reporting three mild symptoms on the instrument.

Symptom scores in our study are lower than naturally-acquired, medically-attended influenza illness (3, 26, 30). Among 221 patients with laboratory-confirmed influenza illness seeking care in ambulatory clinics, peak total symptom scores were 1.6 (equivalent to 51 points on the 32-item FLU-PRO scale), with more predominant chest/respiratory symptoms than we identified in the CHIM study (40). The influenza virus infections that we observed in the CHIM study may be more consistent with asymptomatic or pauci-symptomatic influenza virus infections that are common in community settings (41, 42). For example, in a South African community study with routine, prospective testing for influenza virus infection, approximately 60% of adults aged 19 through 44 years with influenza detections were asymptomatic (41).

There are several strengths to this study. In closely monitored environments using standardized procedures and a GMP challenge virus, our multicenter influenza virus CHIM study was conducted safely and yielded consistent, high attack rates of MMID. Our participant population was diverse and had nearly equal numbers with and without baseline HAI titers ≥40. We corroborated previous observations about the role of HAI in protection from MMID and in modifying viral shedding (25). We also assessed an alternative clinical outcome definition of MMID-2, which was more stringent and more strongly associated baseline with HAI and MN.

There were also limitations to our approach. We included only healthy adult participants, as is standard for CHIM studies, limiting the generalizability of our findings to children, older adults, and persons with chronic medical conditions. Further limiting the generalizability of influenza viral CHIM studies is the nature of inoculation, which may not recapitulate how persons become infected in the community and may contribute to the mild illnesses observed. The challenge virus was closely related to the influenza A (H1N1)pdm09 virus which had circulated for ten years by the time of our study. While about 50% of our participants did not have baseline HAI titers ≥40, it is possible that most had been exposed previously to (H1N1)pdm09 and developed some protective immunity. Our analyses defined a baseline HAI titer ≥40 to be seroprotective; however, this HAI titer is a relative correlate of protection, with previous studies indicating only 50-70% protection from influenza (38). Our use of highly sensitive RT-PCR laboratory diagnostics did not allow us to determine whether the assay was detecting viable virus or nonviable, residual RNA from the challenge inoculum. The more stringent MMID-2 outcome definition, in which we required two or more positive RT-PCR tests, may be more likely to identify participants with true infection. The inclusion of double-blind placebo inoculations could have decreased bias regarding symptom self-reports and clinical assessments. Finally, future influenza vaccine efficacy CHIM trials may seek to assess prevention of influenza illness of greater public health and clinical importance. For influenza vaccines that lessen the severity of illness or have greater efficacy against more severe rather than mild illness, the use of this current challenge model might underestimate the product efficacy.

This trial has advanced NIAID goals to expand the capacity for conducting influenza virus CHIMs. In addition to the results in this report, we collected substantial data and specimens for future exploratory analyses which may help to refine study procedures and better understand the immunology of influenza virus CHIM. Future studies should include more up-to-date influenza viruses, include placebo inoculations to minimize bias in outcome assessment, explore alternative methods to deliver the challenge virus to more closely mimic natural acquisition, and evaluate alternative clinical outcome definitions. There is a robust pipeline of next-generation influenza vaccines and therapeutics in preclinical development (43, 44), and we anticipate that influenza virus CHIMs will help to move them forward to licensure.

## Data Availability

Data to be made available as widely as possible while safeguarding the privacy of participants, and protecting confidential and proprietary data.

## ACKNOWLEDGEMENTS

We thank the members of the data and safety monitoring board for their oversight. From NIAID, we thank Brooke Bozick, Sonja Crandon, Mohamed Elsafy, Binh Hoang, Tena Knudsen, Robin Mason, Kathy Ormanoski, Rhonda Pikaart-Tautges, Christine Oshansky-Weilnau, and Tammy Yokum. From University of Cincinnati, we thank Kristen Buschle, Margie Huron, Jesse LePage, and Monica McNeal. From University of Maryland, we thank Paula Bernal, Melissa Billington, Colleen Boyce, Anna Carmack, Nancy Greenberg, Panagiota Komninou, Hanna LeBuhn, and Jennifer Marron. From Duke University, we thank Jack Anderson, Luis Ballon, Thomas Burke, Kathlene Chmielewski, Min Gao, Thad Gurley, Maria Miggs, and Tim Veldman. From Emory University, we thank Evan Anderson, Colleen Kraft, G. Marshall Lyon, Michele McCullough, and Aneesh Mehta. From Baylor College of Medicine, we thank Connie Rangel. From Emmes, we thank Peter Dubyoski, Alexander Noll, David Santos, Dan Sinnett, Karineh Tarpinian, Julia Weiss, and Shu Yang.

