## Supplement for "A Multi-Center, Controlled Human Infection Study of Influenza A (H1N1)pdm09 in Healthy Adults"

**Influenza Controlled Human Infection Study Group Members (in alphabetical order):**

Azra Blazevic, Kristen Bustle, Wilbur Chen, Lisa Chrisley, Michelle Dickey, Christopher S. Eickhoff, Hana M. El Sahly, Esther Fevrier, Sarah George, Lynn S Harrington, Sonnie Kim, Karen Kotloff, M. Chelsea Lane, Ge Li, Catherine Luke, Karla Mosby, Bradly P. Nicholson, Susan Parker, Rhonda Pikaart-Tautges, Diane J. Post, Marcelo B. Sztein, Janice Tennant, Franklin R. Toapanta, Rachel Tsong

### Supplemental Table 1. Primary and secondary study objectives and endpoints

| Primary Objective | Primary Outcome Measure |
|---|---|
| To evaluate the association of symptomatic RT-PCR-positive influenza virus infection post-challenge and pre-existing HAI antibody titers | Baseline A/Bethesda/MM2/H1N1 hemagglutination inhibition (HAI) antibody GMT association with development of MMID post-challenge, with MMID defined as presence of both of the following assessed through Day 8:<br>1. Viral shedding detected by any approved positive RT-PCR test from NP swab, **and**<br>2. Any one or more of the following symptoms or signs or laboratory findings, as related to the study agent; Arthralgia, Chest tightness, Chills, Conjunctivitis, Nasal congestion, Sinus Congestion, Coryza, Decreased appetite, Diarrhea, Dry Cough, Dyspnea/Shortness of Breath, Fatigue/Tiredness, Fever (>38.0°C), Headache, Lymphopenia (<1000 cells/mL), Myalgia, Nausea, Oxygen Saturation Decrease by ≥3% from baseline, Productive Cough, Rhinorrhea, Sore Throat, and Sweats. |
| **Secondary Objectives** | **Secondary Outcome Measures** |
| To describe viral recovery by quantitative RT-PCR from study subjects at baseline and post-challenge | Frequency, timing, magnitude, and duration of viral shedding in NLF or NP swab collected daily using quantitative RT-PCR from baseline (Day -2 or -1) and daily from Day 2 through Day 8 |
| To describe serum HAI and MN antibody responses post-challenge in healthy subjects by infection status | Baseline and post-challenge HAI and MN antibody GMTs from serum measured at baseline (Day -2) and at Days, 8, 29, and 61 by infection status (RT-PCR-confirmed symptomatic, RT-PCR-confirmed asymptomatic, RT-PCR-negative symptomatic)<br><br>Percentage of healthy subjects achieving HAI and MN seroconversion (defined as either a pre-challenge titer <1:10 and a post-challenge titer ≥1:40 or a pre-challenge titer ≥1:10 and a minimum four-fold rise in post-challenge antibody titer) from serum measured at baseline (Day -2) and on Days 8, 29, and 61 |
| To evaluate the association of asymptomatic RT-PCR-positive influenza virus infection (viral shedding) post-challenge and pre-existing HAI antibody titers | Baseline A/Bethesda/MM2/H1N1 HAI antibody GMT measured at baseline (Day -2) association with asymptomatic viral shedding by RT-PCR through Day 8 |
| To evaluate the association of symptomatic RT-PCR-negative status post-challenge and pre-existing HAI antibody titers | Baseline A/Bethesda/MM2/H1N1 HAI antibody GMT measured at baseline (Day -2) association with development of any Flu-PRO symptoms post-challenge without influenza virus detection through Day 8 |
| To determine the frequency of serious adverse events (SAE) post-challenge | Frequency of serious adverse events (SAE) post-challenge through the inpatient stay |

**Supplemental Table 2. Participant demographic and baseline characteristics, overall and by site**

| | | Saint Louis University (N=17) | | Cincinnati Children's Hospital (N=24) | | University of Maryland (N=20) | | Duke University (N=15) | | All Participants (N=76) | |
|---|---|---|---|---|---|---|---|---|---|---|---|
| Variable | Characteristic | n | % | n | % | n | % | n | % | n | % |
| Sex | Male | 9 | 53 | 17 | 71 | 14 | 70 | 6 | 40 | 46 | 61 |
| | Female | 8 | 47 | 7 | 29 | 6 | 30 | 9 | 60 | 30 | 39 |
| Age | 18-29 | 8 | 47 | 11 | 46 | 5 | 25 | 6 | 40 | 30 | 39 |
| | 30-39 | 5 | 29 | 7 | 29 | 9 | 45 | 3 | 20 | 24 | 32 |
| | 40-49 | 4 | 24 | 6 | 25 | 6 | 30 | 6 | 40 | 22 | 29 |
| BMI | <30 | 15 | 88 | 18 | 75 | 8 | 40 | 9 | 60 | 50 | 66 |
| | ≥30 | 2 | 12 | 6 | 25 | 12 | 60 | 6 | 40 | 26 | 34 |
| Ethnicity | Not Hispanic or Latino | 16 | 94 | 24 | 100 | 17 | 85 | 14 | 93 | 71 | 93 |
| | Hispanic or Latino | 1 | 6 | - | - | 3 | 15 | 1 | 7 | 5 | 7 |
| | Not Reported | - | - | - | - | - | - | - | - | - | - |
| | Unknown | - | - | - | - | - | - | - | - | - | - |
| Race | American Indian or Alaska Native | - | - | - | - | 1 | 5 | - | - | 1 | 1 |
| | Asian | - | - | - | - | 1 | 5 | - | - | 1 | 1 |
| | Native Hawaiian or Other Pacific Islander | - | - | - | - | - | - | - | - | - | - |
| | Black or African American | 2 | 12 | 11 | 46 | 14 | 70 | 8 | 53 | 35 | 46 |
| | White | 13 | 76 | 13 | 54 | 3 | 15 | 5 | 33 | 34 | 45 |
| | Multi-Racial | 2 | 12 | - | - | - | - | 1 | 7 | 3 | 4 |
| | Unknown | - | - | - | - | 1 | 5 | 1 | 7 | 2 | 3 |

### Supplemental Table 3. Hemagglutination inhibition endpoints by study day and infection status

| Study Day | Statistic | RT-PCR Positive Symptomatic (MMID-1) | At Least Twice RT- PCR Positive Symptomatic (MMID-2) | RT-PCR Positive Asymptomatic | RT-PCR Negative (None Positive) | RT-PCR Positive (One or More) | RT-PCR Positive (Two or More) |
|---|---|---|---|---|---|---|---|
| Baseline | n | 54 | 38 | 8 | 14 | 62 | 43 |
| | GMT (95% CI)b | 36.8 (26.1, 52.0) | 27.6 (18.4,41.4) | 20.0 (6.9, 58.0) | 117.9 (61.3,227.0) | 34.0 (24.7,47.0) | 26.3 (18.0,38.6) |
| | Seroprotection % (95% CI)c | 44.4 (30.9,58.6) | 34.2 (19.6,51.4) | 25 (3.2, 65.1) | 78.6 (49.2,95.3) | 41.9 (29.5,55.2) | 32.6 (19.1,48.5) |
| Day 8 | n | 53 | 37 | 8 | 14 | 61 | 42 |
| | GMT (95% CI) | 46.1 (33.2,64.0) | 34.6 (23.1,52.0) | 23.7 (8.9, 63.1) | 122.5 (66.9,224.6) | 42.2 (31.1,57.5) | 31.9 (21.8,46.8) |
| | GMFR (95% CI)b | 1.2 (1.1, 1.5) | 1.3 (1.0, 1.6) | 1.2 (0.8, 1.7) | 1.0 (0.8, 1.3) | 1.2 (1.1, 1.4) | 1.2 (1.0, 1.5) |
| | Seroprotection % (95% CI) | 60.4 (46, 73.5) | 48.6 (31.9,65.6) | 25 (3.2, 65.1) | 85.7 (57.2,98.2) | 55.7 (42.4,68.5) | 45.2 (29.8,61.3) |
| | Seroconversion % (95% CI)c | 3.8 (0.5, 13) | 5.4 (0.7, 18.2) | 0 (0, 36.9) | 0 (0, 23.2) | 3.3 (0.4, 11.3) | 4.8 (0.6, 16.2) |
| Day 29 | n | 46 | 31 | 7 | 14 | 53 | 35 |
| | GMT (95% CI) | 87.4 (65.7,116.2) | 74.8 (51.1,109.6) | 63.9 (29.2,139.9) | 126.6 (72.3,221.9) | 83.8 (64.6,108.8) | 73.5 (51.9,103.9) |
| | GMFR (95% CI) | 2.3 (1.6, 3.1) | 2.6 (1.7, 4.0) | 2.7 (1.0, 7.5) | 1.1 (0.9, 1.3) | 2.3 (1.7, 3.1) | 2.6 (1.8, 3.9) |
| | Seroprotection % (95% CI) | 82.6 (68.6,92.2) | 77.4 (58.9,90.4) | 71.4 (29, 96.3) | 92.9 (66.1,99.8) | 81.1 (68, 90.6) | 77.1 (59.9,89.6) |
| | Seroconversion % (95% CI) | 28.3 (16, 43.5) | 32.3 (16.7,51.4) | 28.6 (3.7, 71) | 0 (0, 23.2) | 28.3 (16.8,42.3) | 31.4 (16.9,49.3) |
| Day 61 | n | 43 | 29 | 6 | 13 | 49 | 33 |
| | GMT (95% CI) | 78.0 (55.5,109.7) | 65.5 (41.8,102.7) | 71.3 (33.6,151.1) | 123.1 (63.9,237.3) | 77.2 (56.9,104.7) | 65.0 (43.4,97.3) |
| | GMFR (95% CI) | 2.0 (1.5, 2.7) | 2.3 (1.5, 3.5) | 2.6 (1.1, 6.3) | 1.1 (0.8, 1.4) | 2.1 (1.6, 2.7) | 2.4 (1.6, 3.4) |
| | Seroprotection % (95% CI) | 76.7 (61.4,88.2) | 72.4 (52.8,87.3) | 83.3 (35.9, 99.6) | 92.3 (64, 99.8) | 77.6 (63.4,88.2) | 72.7 (54.5,86.7) |
| | Seroconversion % (95% CI) | 18.6 (8.4, 33.4) | 24.1 (10.3,43.5) | 33.3 (4.3, 77.7) | 0 (0, 24.7) | 20.4 (10.2,34.3) | 24.2 (11.1,42.3) |

Notes: n = Number of participants in the Modified Intent-to-Treat population with available results; GMT = Geometric Mean Titer; GMFR = Geometric Mean Fold Rise.

[a] If ever reported during the challenge period (Days 2 to 8).

[b] Confidence Interval calculated based on the Student's T distribution.

[c] Exact binomial confidence interval calculated using the Clopper-Pearson methodology.

### Supplemental Table 4. Microneutralization endpoints by study day and infection status

| Study Day | Statistic | RT-PCR Positive Symptomatic (MMID-1) | At Least Twice RT- PCR Positive Symptomatic (MMID-2) | RT-PCR Positive Asymptomatic | RT-PCR Negative (None Positive) | RT-PCR Positive (One or More) | RT-PCR Positive (Two or More) |
|---|---|---|---|---|---|---|---|
| Baseline | n | 54 | 38 | 8 | 14 | 62 | 43 |
| | GMT (95% CI)b | 62.9 (43.1,91.9) | 45.1 (29.3, 69.5) | 44.3 (13.8, 142.6) | 276.7 (124.8, 613.1) | 60.1 (42.3, 85.4) | 45.2 (30.0, 68.2) |
| | Seroprotection % (95% CI)c | 66.7 (52.5, 78.9) | 60.5 (43.4, 76) | 50 (15.7, 84.3) | 92.9 (66.1, 99.8) | 64.5 (51.3, 76.3) | 60.5 (44.4, 75) |
| Day 8 | n | 53 | 37 | 8 | 14 | 61 | 42 |
| | GMT (95% CI) | 87.7 (61.7, 124.6) | 66.8 (43.9, 101.6) | 52.4 (16.6, 165.5) | 250.6 (112.8, 556.9) | 81.9 (58.9, 114.0) | 64.0 (42.8, 95.7) |
| | GMFR (95% CI)b | 1.4 (1.2, 1.7) | 1.5 (1.2, 1.9) | 1.2 (0.8, 1.8) | 0.9 (0.8, 1.0) | 1.4 (1.2, 1.6) | 1.4 (1.1, 1.8) |
| | Seroprotection % (95% CI) | 77.4 (63.8, 87.7) | 73 (55.9, 86.2) | 62.5 (24.5, 91.5) | 92.9 (66.1, 99.8) | 75.4 (62.7, 85.5) | 71.4 (55.4, 84.3) |
| | Seroconversion % (95% CI)c | 7.5 (2.1, 18.2) | 10.8 (3, 25.4) | 0 (0, 36.9) | 0 (0, 23.2) | 6.6 (1.8, 15.9) | 9.5 (2.7, 22.6) |
| Day 29 | n | 46 | 31 | 7 | 14 | 53 | 35 |
| | GMT (95% CI) | 159.5 (115.7, 220.1) | 131.6 (87.5, 197.7) | 103.1 (41.5, 255.9) | 320.0 (157.9, 648.5) | 150.6 (112.1, 202.4) | 126.9 (86.9, 185.5) |
| | GMFR (95% CI) | 2.5 (1.8, 3.4) | 3.0 (2.0, 4.5) | 2.8 (1.3, 6.2) | 1.2 (1.0, 1.3) | 2.5 (1.9, 3.3) | 3.0 (2.1, 4.3) |
| | Seroprotection % (95% CI) | 89.1 (76.4, 96.4) | 87.1 (70.2, 96.4) | 85.7 (42.1, 99.6) | 100 (76.8, 100) | 88.7 (77, 95.7) | 85.7 (69.7, 95.2) |
| | Seroconversion % (95% CI) | 26.1 (14.3, 41.1) | 32.3 (16.7, 51.4) | 28.6 (3.7, 71) | 0 (0, 23.2) | 26.4 (15.3, 40.3) | 31.4 (16.9, 49.3) |
| Day 61 | n | 43 | 29 | 6 | 13 | 49 | 33 |
| | GMT (95% CI) | 151.8 (108.7, 211.9) | 126.5 (84.9, 188.2) | 107.2 (30.4, 377.8) | 287.6 (134.9, 613.2) | 145.4 (106.3, 199.0) | 121.0 (82.3, 177.9) |
| | GMFR (95% CI) | 2.2 (1.7, 2.9) | 2.8 (1.9, 4.1) | 2.6 (1.1, 6.6) | 1.1 (0.9, 1.3) | 2.2 (1.7, 2.9) | 2.8 (2.0, 3.9) |
| | Seroprotection % (95% CI) | 90.7 (77.9, 97.4) | 89.7 (72.6, 97.8) | 83.3 (35.9, 99.6) | 100 (75.3, 100) | 89.8 (77.8, 96.6) | 87.9 (71.8, 96.6) |
| | Seroconversion % (95% CI) | 18.6 (8.4, 33.4) | 27.6 (12.7, 47.2) | 50 (11.8, 88.2) | 0 (0, 24.7) | 22.4 (11.8, 36.6) | 30.3 (15.6, 48.7) |

Notes: n = Number of participants in the Modified Intent-to-Treat population with available results; GMT = Geometric Mean Titer; GMFR = Geometric Mean Fold Rise.

[a] If ever reported during the challenge period (Days 2 to 8).

[b] Confidence Interval calculated based on the Student's T distribution.

[c] Exact binomial confidence interval calculated using the Clopper-Pearson methodology.

### Supplemental Table 5. Neuraminidase inhibition endpoints by study day and infection status

| Study Day | Statistic | RT-PCR Positive Symptomatic (MMID-1) | At Least Twice RT- PCR Positive Symptomatic (MMID-2) | RT-PCR Positive Asymptomatic | RT-PCR Negative (None Positive) | RT-PCR Positive (One or More) | RT-PCR Positive (Two or More) |
|---|---|---|---|---|---|---|---|
| Baseline | n | 54 | 38 | 8 | 14 | 62 | 43 |
| | GMT (95% CI)b | 584.6 (424.7,804.7) | 506.1 (345.4,741.6) | 863.3 (358.3,2080.3) | 844.2 (508.1,1402.7) | 614.8 (458.4,824.4) | 565.4 (396.0,807.2) |
| Day 8 | n | 53 | 37 | 8 | 14 | 61 | 42 |
| | GMT (95% CI) | 692.2 (516.1,928.4) | 604.6 (424.4,861.1) | 1031.5 (521.3,2041.0) | 875.9 (531.8,1442.5) | 729.4 (559.0,951.7) | 670.3 (483.7,928.9) |
| | GMFR (95% CI)b | 1.2 (1.1, 1.3) | 1.2 (1.1, 1.3) | 1.2 (0.9, 1.5) | 1.0 (0.9, 1.2) | 1.2 (1.1, 1.3) | 1.2 (1.1, 1.3) |
| | Seroconversion % (95% CI)c | 0 (0, 6.7) | 0 (0, 9.5) | 0 (0, 36.9) | 0 (0, 23.2) | 0 (0, 5.9) | 0 (0, 8.4) |
| Day 29 | n | 46 | 31 | 7 | 14 | 53 | 35 |
| | GMT (95% CI) | 979.4 (770.9,1244.4) | 952.8 (711.8,1275.4) | 1707.1 (984.1,2961.2) | 1176.6 (795.3,1740.6) | 1054.0 (845.5,1314.0) | 1043.5 (788.3,1381.3) |
| | GMFR (95% CI) | 1.7 (1.5, 2.1) | 2.0 (1.6, 2.4) | 2.2 (1.1, 4.3) | 1.4 (1.1, 1.7) | 1.8 (1.5, 2.1) | 1.9 (1.6, 2.4) |
| | Seroconversion % (95% CI)c | 10.9 (3.6, 23.6) | 16.1 (5.5, 33.7) | 14.3 (0.4, 57.9) | 0 (0, 23.2) | 11.3 (4.3, 23) | 14.3 (4.8, 30.3) |
| Day 61 | n | 43 | 29 | 6 | 13 | 49 | 33 |
| | GMT (95% CI) | 1052.0 (818.4,1352.3) | 1029.2 (753.5,1405.8) | 1629.3 (884.5,3001.0) | 1111.5 (681.1,1814.0) | 1109.9 (882.2,1396.3) | 1101.7 (826.1,1469.2) |
| | GMFR (95% CI) | 2.0 (1.6, 2.3) | 2.2 (1.8, 2.8) | 2.0 (0.9, 4.2) | 1.3 (1.1, 1.6) | 2.0 (1.7, 2.3) | 2.1 (1.7, 2.6) |
| | Seroconversion % (95% CI)c | 14 (5.3, 27.9) | 20.7 (8, 39.7) | 16.7 (0.4, 64.1) | 0 (0, 24.7) | 14.3 (5.9, 27.2) | 18.2 (7, 35.5) |

Notes: n = Number of participants in the Modified Intent-to-Treat population with available results; GMT = Geometric Mean Titer; GMFR = Geometric Mean Fold Rise.

[a] If ever reported during the challenge period (Days 2 to 8).

[b] Confidence Interval calculated based on the Student's t- distribution.

[c] Exact binomial confidence interval calculated using the Clopper-Pearson methodology.

### Supplemental Table 6. Safety outcomes

| | All Participants (N=76) | |
|---|---|---|
| **Participants with** | **n** | **%** |
| At least one unsolicited adverse event | 46 | 61 |
| At least one related unsolicited adverse event | 26 | 34 |
| Mild (Grade 1) | 24 | 32 |
| Moderate (Grade 2) | 2 | 3 |
| Severe (Grade 3) | - | - |
| At least one severe (Grade 3) unsolicited adverse event | - | - |
| Related | - | - |
| Unrelated | - | - |
| At least one serious adverse event | - | - |
| At least one related, serious adverse event | - | - |
| At least one adverse event leading to early termination[b] | - | - |
| At least one hematology laboratory adverse event[c] | 13 | 17 |
| Mild (Grade 1) | 13 | 17 |
| Moderate (Grade 2) | - | - |
| Severe (Grade 3) | - | - |
| At least one chemistry laboratory adverse event[c] | 7 | 9 |
| Mild (Grade 1) | 7 | 9 |
| Moderate (Grade 2) | - | - |
| Severe (Grade 3) | - | - |

Notes: N = Number of participants in the Safety population.

a Participants are counted once for each category regardless of the number of events, and only a participant's maximum severity is counted.

b As reported on the Adverse Event eCRF.

c Only findings that are not part of Mild-to-Moderate Influenza Disease and occurred post-challenge are considered.

**Supplemental Figure 1. Frequency of positive viral shedding results by study day, safety population**

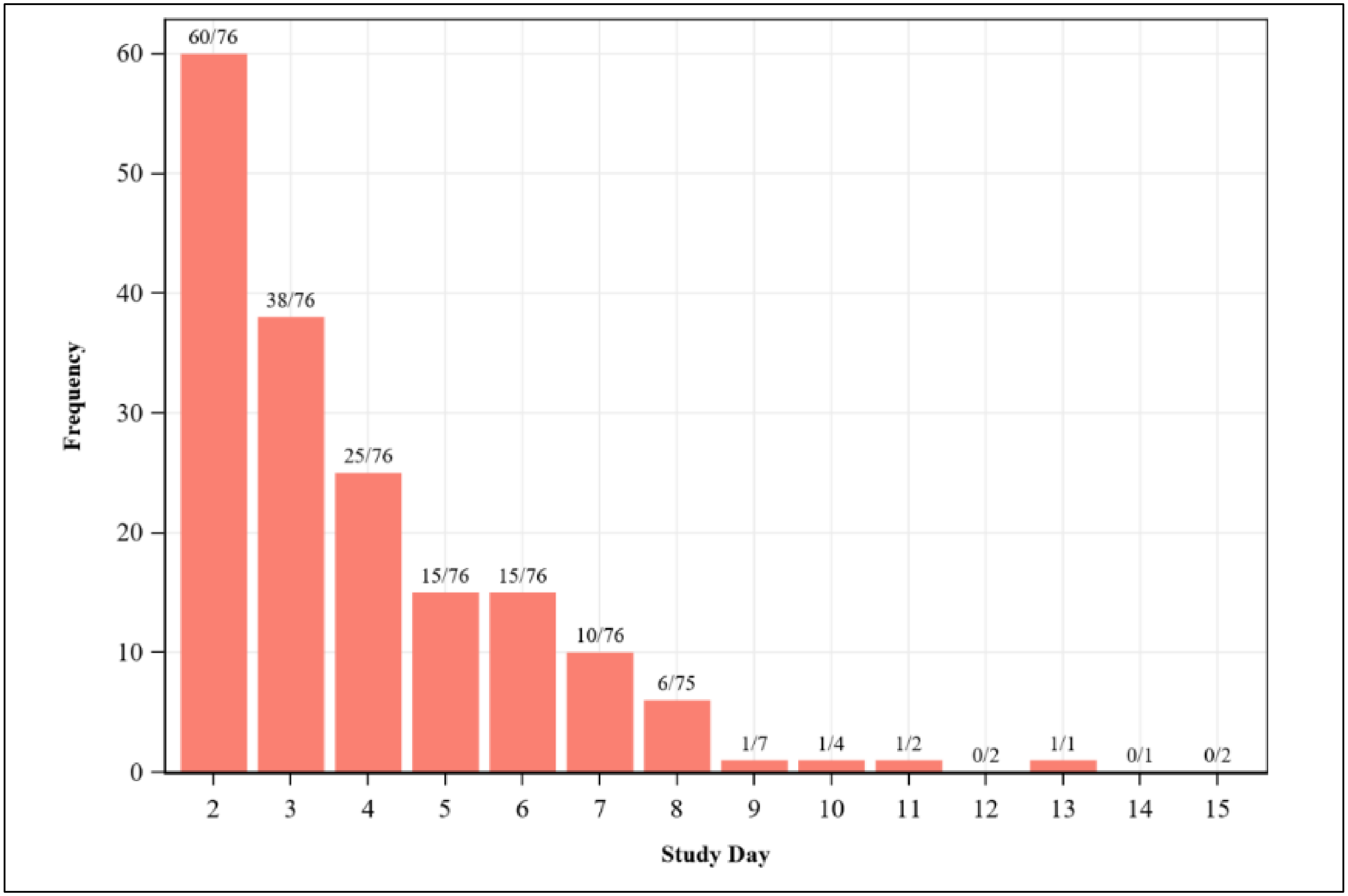

**Supplemental Figure 2. Predicted probabilities of MMID according to baseline HAI against challenge virus, univariable model, modified intent-to-Treat Population**

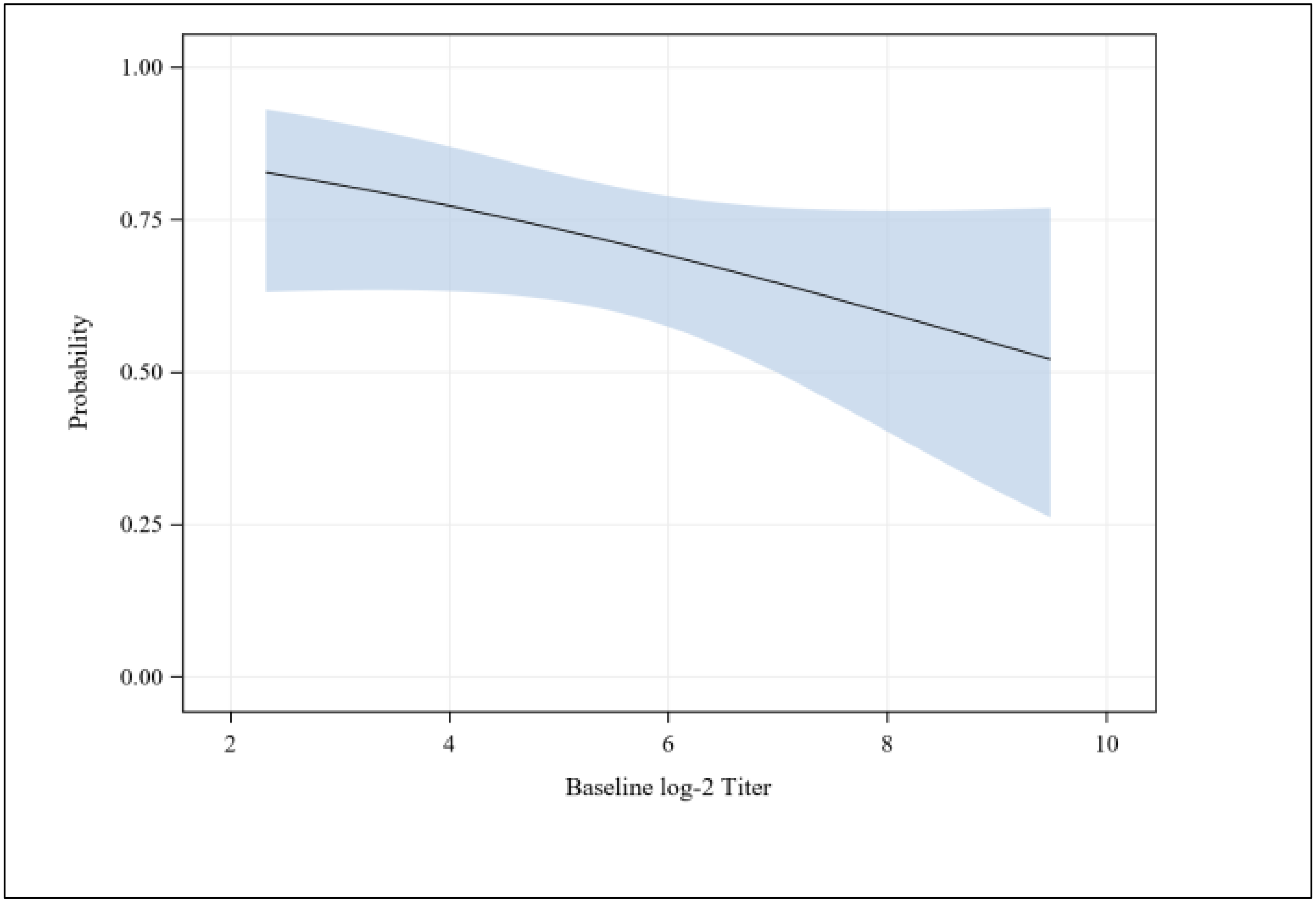

**Supplemental Figure 3. Predicted probabilities of MMID according to baseline MN against challenge virus, univariable model, modified intent-to-Treat Population**

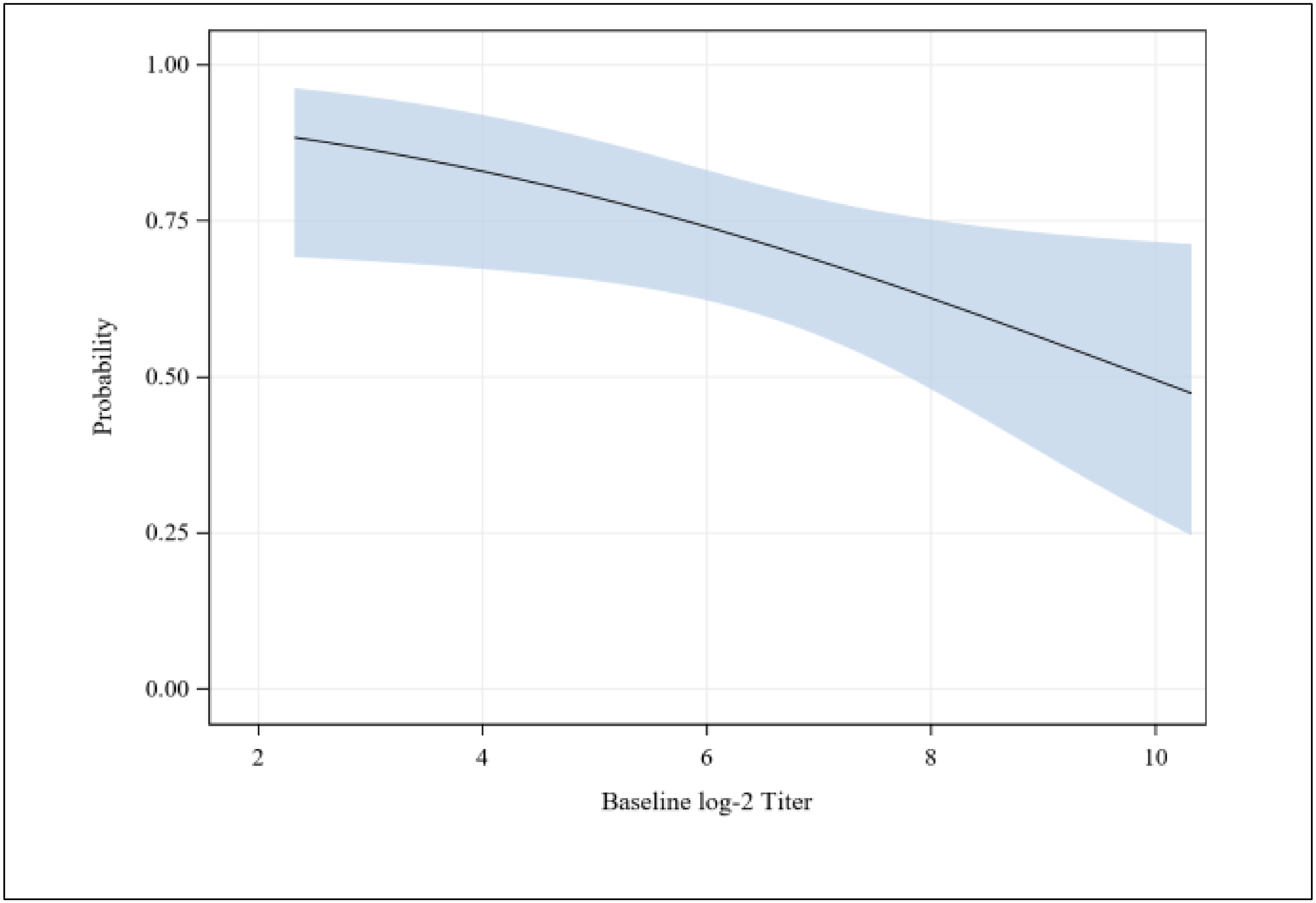

**Supplemental Figure 4. Predicted probabilities of MMID according to baseline NAI against challenge virus, univariable model, modified Intent-to-Treat Population**

¶

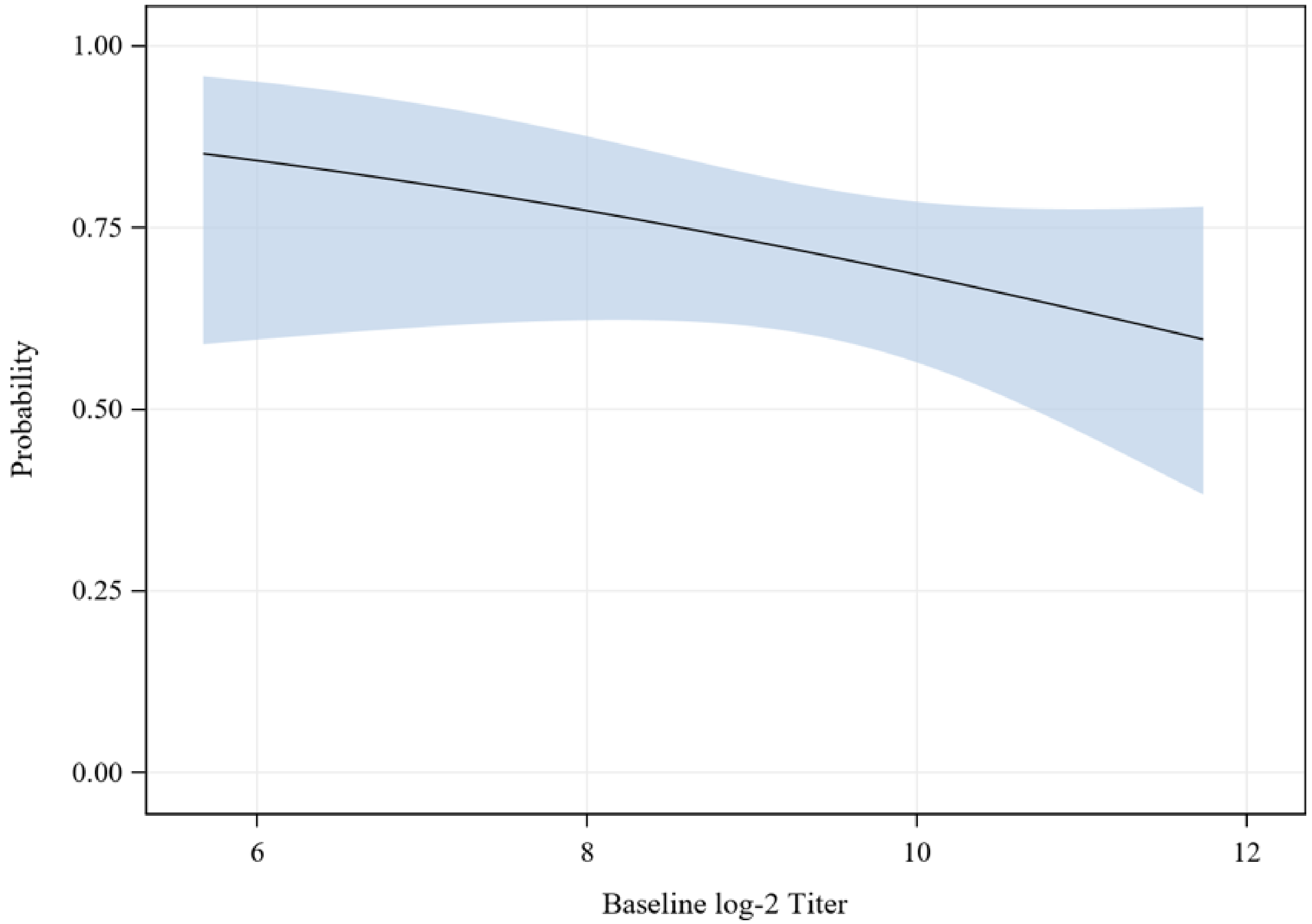


¶
¶

**Supplemental Figure 5. Kaplan-Meier curves for time to viral shedding by baseline HAI seroprotection status, modified Intent-to-Treat Population**

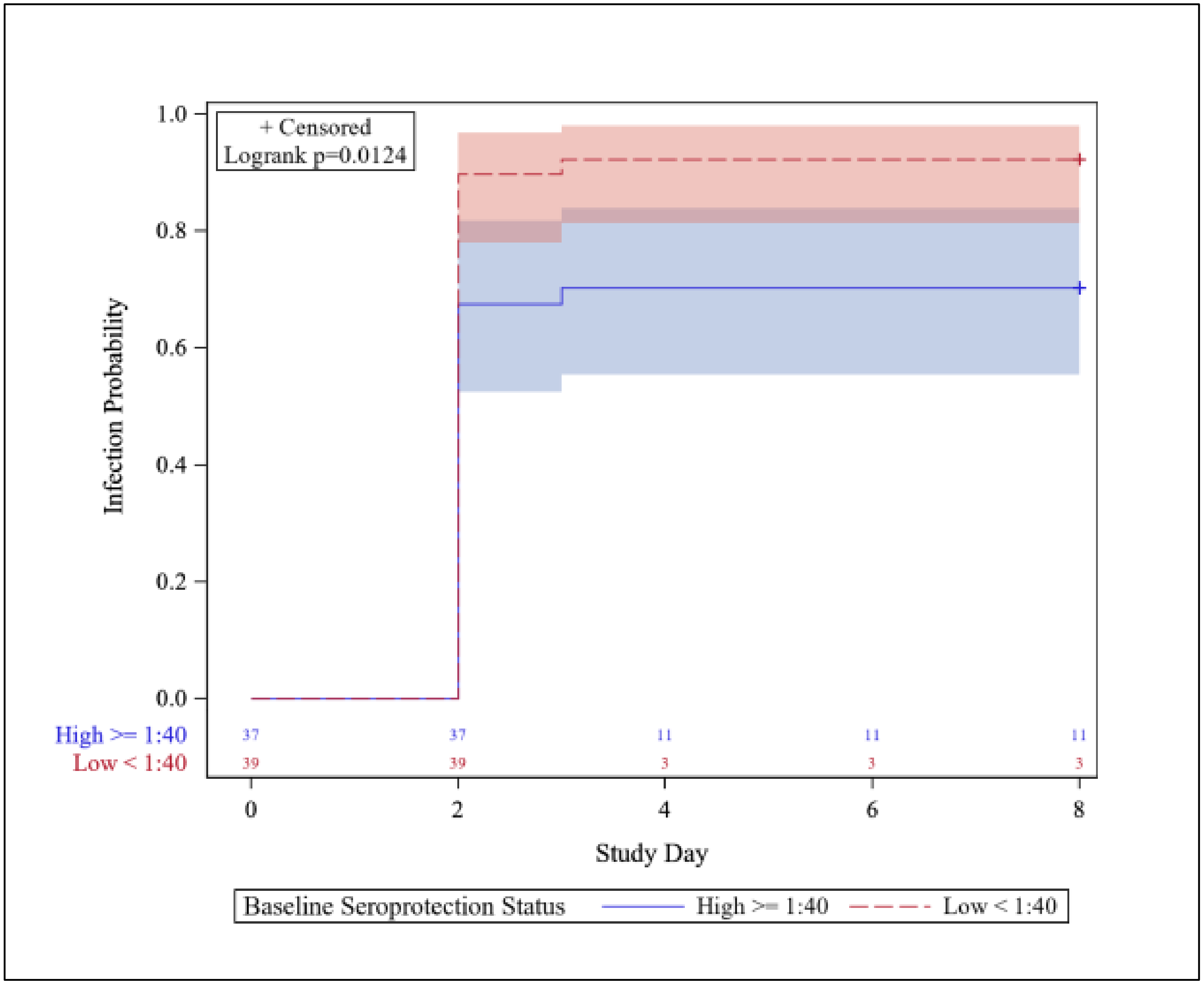

**Supplemental Figure 6. Kaplan-Meier curves for time to viral shedding by baseline MN seroprotection status, modified Intent-to-Treat Population**

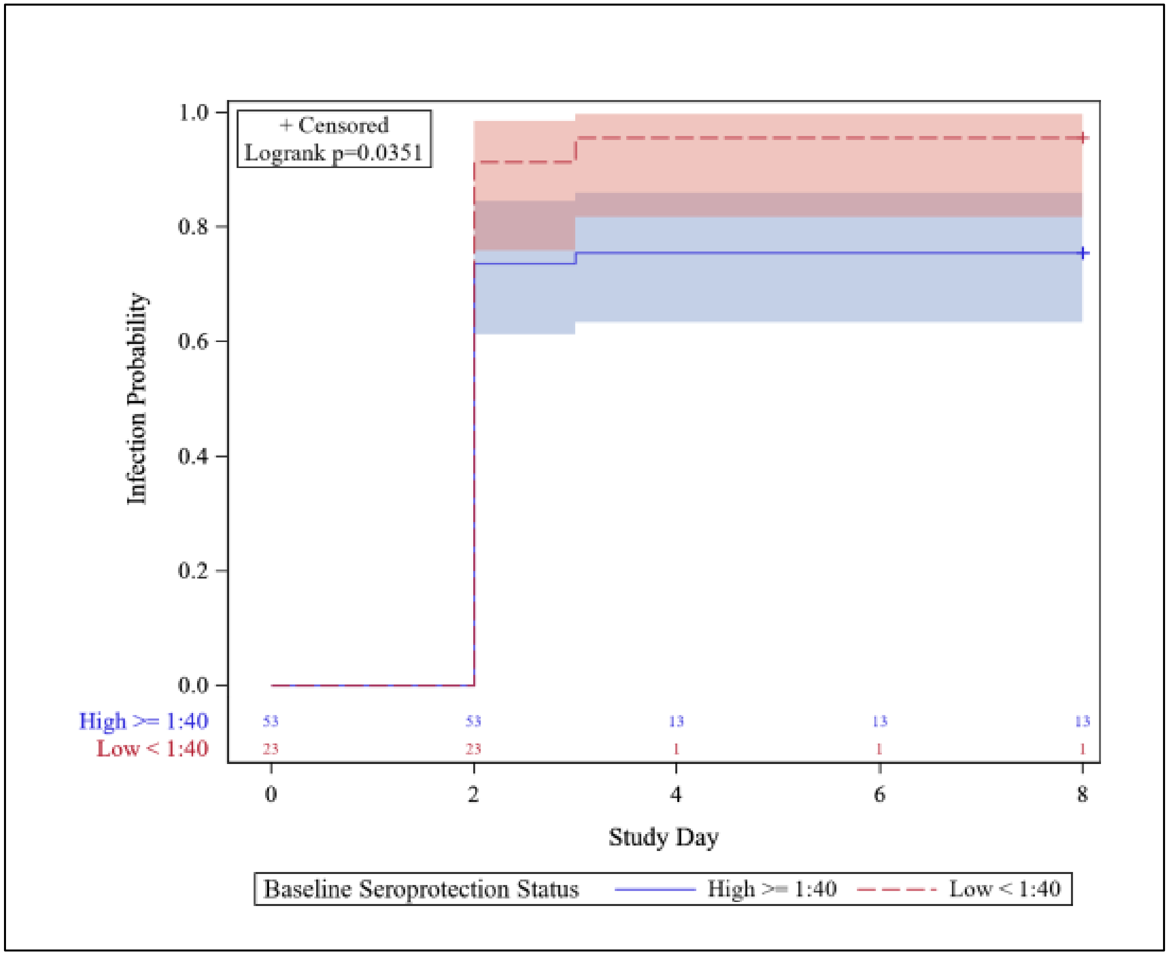

**Supplemental Figure 7. Hemagglutination inhibition, microneutralization, and neuraminidase inhibition antibody geometric mean titers, by infection status (using MMID-2) and study day**

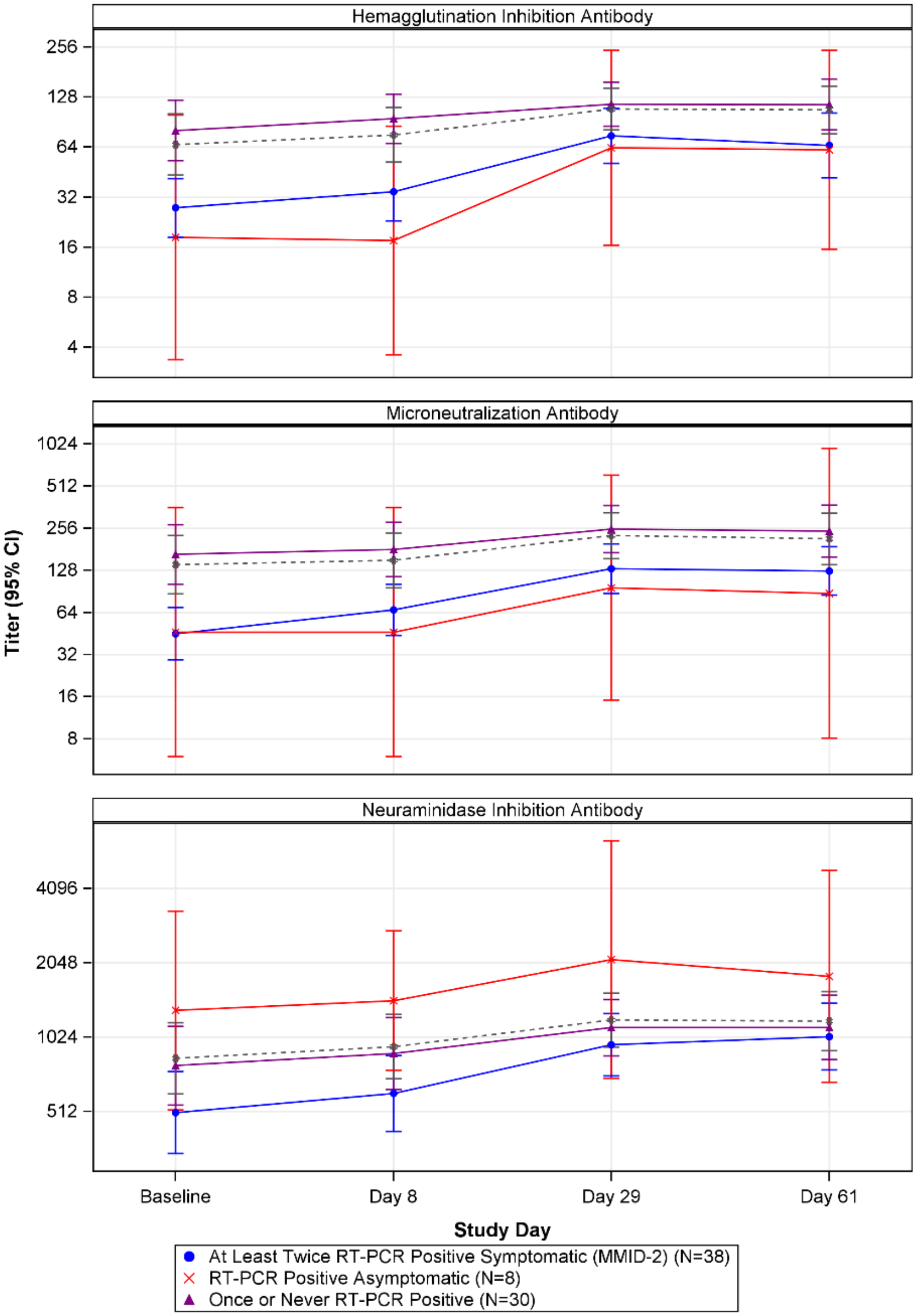


Note: The gray dashed line displays the trajectory of the combined RT-PCR positive asymptomatic and the once or never RT-PCR positive groups (i.e. participants who did not meet the definition of MMID-2).
